# Short- and long-term effects of imatinib in patients hospitalised for COVID-19 infection: A randomised controlled trial

**DOI:** 10.1101/2024.02.02.24302227

**Authors:** Alex L. E. Halme, Sanna Laakkonen, Jarno Rutanen, Olli P. O. Nevalainen, Marjatta Sinisalo, Saana Horstia, Jussi M. J. Mustonen, Negar Pourjamal, Aija Vanhanen, Solidarity Finland Investigators, Tuomas Rosberg, Andreas Renner, Markus Perola, Erja-Leena Paukkeri, Riitta-Liisa Patovirta, Seppo Parkkila, Juuso Paajanen, Taina Nykänen, Jarkko Mäntylä, Marjukka Myllärniemi, Tiina Mattila, Maarit Leinonen, Alvar Külmäsu, Pauliina Kuutti, Ilari Kuitunen, Hanna-Riikka Kreivi, Tuomas P. Kilpeläinen, Heikki Kauma, Ilkka E. J. Kalliala, Petrus Järvinen, Riina Hankkio, Taina Hammarén, Thijs Feuth, Hanna Ansakorpi, Riikka Ala-Karvia, Gordon H. Guyatt, Kari A. O. Tikkinen

**Author notes:** **Corresponding author:** Professor Kari A. O. Tikkinen, Faculty of Medicine, University of Helsinki, Biomedicum 2 B, P.O. Box 13, Tukholmankatu 8 B, 00290 Helsinki, Finland. **Collaborators (Solidarity Finland Investigators):** Tero Ala-Kokko, Maarit Gockel, Susanna Haapanen, Mia Haukipää, Sanna Lehtonen, Eeva-Maija Nieminen, Joni Niskanen, Jarmo Oksi, Ulla Otava, Taija Rutanen, Mari Saalasti, Petri Salmela, Joni Savolainen, Susanna Tuominen, Henri Vartiainen, Heli Ylä-Outinen.

## Abstract

We report the short- and long-term results of the SOLIDARITY Finland on mortality and other patient-important outcomes in patients hospitalised for COVID-19. Between 08/2021 and 03/2023, we randomised 156 patients in 15 hospitals. In the imatinib group, 7.2% of patients had died at 30 days and 13.3% at 1 year and in the standard of care group 4.1% and 8.3% (adjusted HR at 30 days 1.09, 95% CI 0.23–5.07). In a meta-analysis of randomised trials of imatinib versus standard of care (n=732), allocation to imatinib was associated with a mortality risk ratio of 0.73 (95% CI 0.32–1.63). At 1-year, self-reported recovery occurred in 79.0% in imatinib and in 88.3% in standard of care (RR 0.91, 95% CI 0.78-1.06). Of the 21 potential long COVID symptoms, patients often reported moderate or major bother from fatigue (24%), sleeping problems (19%) and memory difficulties (17%). We found no convincing difference between imatinib and standard of care groups in quality of life or symptom outcomes. The evidence raises serious doubts regarding the benefit of imatinib in reducing mortality, improving recovery, and preventing potential long COVID symptoms when given to patients hospitalised for COVID-19.

## Introduction

The coronavirus disease 2019 (COVID-19) pandemic has been the largest public health crisis of the last century. While currently patients with SARS-CoV-2 infection are overwhelmingly asymptomatic or experience only mild to moderate symptoms, early in the pandemic the infection was often much more severe resulting in significant morbidity and mortality.^1–4^ As of January 2024, more than seven million COVID-19 deaths have been reported to the World Health Organization (WHO).^5^

Since the inception of the pandemic, numerous trials have attempted to identify potential treatments, mainly antiviral or immunomodulatory drugs.^6–12^ Several medications, including dexamethasone, remdesivir, baricitinib and tocilizumab, decrease mortality in patients hospitalized with COVID-19.^6–11^ As none of the beforementioned drugs shown to be effective for hospitalized COVID-19 patients have had large effects, we need to find new therapies.

Preclinical studies suggested that imatinib, a small molecule tyrosine kinase inhibitor, improves endothelial barrier integrity and reverses pulmonary oedema in acute lung injury.^13,14^ A placebo-controlled trial randomized 400 COVID-19 patients requiring supplemental oxygen administration to imatinib vs placebo. This trial, published in 2021, reported that 15 (8%) patients receiving imatinib and 27 (14%) patients receiving placebo had died at 28-day follow-up (HR 0.51, 95% CI 0.27–0.95).^15^ However, subsequent small trials have been inconclusive.^16,17^

Since August 2021, the Solidarity Finland trial, in collaboration with the WHO, has evaluated the benefits and harms of imatinib compared to local standard of care in patients hospitalized for COVID-19. Our study is the first to investigate long-term effects of imatinib treatment on COVID-19 and among the few COVID-19 trials on any drug treatment to include a long-term follow-up on overall quality of life and long COVID (synonymously post COVID-19 condition) symptoms.^18,19^ Here, we first report the results of our trial during the initial hospitalisation phase including the effect of imatinib treatment on hospital mortality. Second, we report the results of 12-month follow-up including mortality, overall recovery, quality of life and other patient-important outcomes typically associated with long COVID.

## Methods

### Study design and participants

We conducted the randomized, pragmatic, multicentre, parallel, open-label Solidarity Finland trial (EudraCT 2020-001784-88; clinicaltrials.gov NCT05220280) in 15 hospitals in Finland with patient recruitment from August 2021 to March 2023. We randomized adult patients hospitalized for PCR-confirmed COVID-19 to receive either local standard of care or local standard of care with orally administered imatinib.

Patients eligible for inclusion were i) at least 18 years old, ii) had a laboratory-confirmed SARS-CoV-2 infection, iii) were hospitalized, iv) the patient or their next of kin provided informed consent, and v) no planned transfer of the patient to a hospital not participating in the trial within the next 72 hours. We advised clinicians to recruit patients to the trial only if patients were symptomatic of COVID-19 (not if they were COVID-19 positive but asymptomatic of COVID-19 and hospitalized for other reasons).

We excluded patients from the Solidarity Finland trial if they: i) had a severe co-morbidity with life expectancy <3 months according to investigators assessment, ii) had high AST/ALT (5 times above upper normal limit), iii) had an eGFR < 30 ml/min, iv) had other acute diseases within the previous 7 days (such as myocardial infarction or unstable angina pectoris), v) were pregnant or breastfeeding, vi) had liver cirrhosis or hepatitis B, vii) were already participating in another trial, or viii) were for any other reason unable to participate in the trial (Supplementary Appendix 1).

The trial complied with all relevant ethical regulations and received approval by the Finnish Medicines Agency Fimea (33/2020), National Committee on Medical Research Ethics TUKIJA (56/2021) and ethics board of the Helsinki University Hospital HUS (1866/2021). Additionally, the trial received approval locally from each of the 15 participating hospitals. In reporting the results of this trial, we adhered to CONSORT guidelines with relevant extensions.^20–22^

### Randomisation and masking

We used online Castor EDC software (https://www.castoredc.com) to randomize patients and collect data. Before central concealed treatment assignment, the local investigator inputted which study drugs are available at their hospital (imatinib, infliximab or both). Patients could only be randomized to receive the drug if they had no contraindications (Supplementary Appendix 1) and if the drug was available at the hospital at which they were treated (imatinib was available in all Solidarity Finland hospitals during the whole trial; infliximab was available in most hospitals most of the time). We allocated patients equally to all available study arms available to them. In this article, we report the patients who participated in the trial of imatinib vs standard of care as well as the patients who participated in the trial between three arms (standard of care, imatinib and infliximab) and were randomised to either imatinib or standard of care.

### Procedures and outcomes

Patients in the imatinib group received 400 mg of oral imatinib from randomisation until discharge or 14 days in hospital.

#### Short-term follow-up

We recorded overall mortality at 30 days, length of hospital stay and need of respiratory support during hospital phase with an electronic case report form. During the hospital phase, we also collected information on serious adverse events (SAE) or suspected unexpected serious adverse reactions (SUSAR).

#### Long-term follow-up (1 year follow-up from hospital admission)

We followed patients for 1 year and collected: i) mortality up to January 2024 from the Digital and Population Data Services Agency (Helsinki, Finland) and ii) a survey addressing patient-reported outcomes (Supplementary Appendix 2). Our multidisciplinary team of clinicians, methodologists, and patient partners (TR, JS) participated in developing the survey questionnaire to assess long-term recovery and symptoms typically associated with post-COVID syndrome, synonymously long COVID (Supplementary Appendix 3). We used the modified Medical Research Council dyspnoea scale to assess exertional dyspnoea, and the EQ-5D-5L and the visual analogue scale (VAS) scale to measure mobility, self-care, usual daily activities, general pain/discomfort, anxiety/depression, and an overall impression of health. In addition to Finnish-language, we translated questionnaires, consent forms and information leaflets from Finnish to Albanian, Arabic, English, Estonian, Persian, Russian, Somalian, and Swedish for patients speaking those languages.

### Statistical analysis

All statistical analyses were intention-to-treat analyses with two-sided p-values and a significance level of 5%. Following the WHO SOLIDARITY trial protocol,^23^ we did not prespecify sample size. We report baseline characteristics as median or mean values and interquartile ranges (IQR) or standard deviations. We used Kaplan-Meier survival curves and Cox regression analysis to compare survival of patients over time and tested the proportional hazard assumption with Schoenfeld residuals. We compared overall survival at 30 days and 1 year. We calculated the difference between groups as both an unadjusted and adjusted hazard ratio (HR) for possibly prognostic baseline imbalances (adjusted for COVID-19 severity at admission, age stratified in decades and sex) alongside their associated 95% confidence intervals (CI). We reported the adjusted HRs in the main text because adjusted and unadjusted HRs were similar. We used Fisher’s exact test to compare categorical outcomes, the Mann–Whitney U test for continuous outcomes, and Welch’s t-test to calculate confidence intervals for absolute differences of continuous outcomes. We conducted all statistical analyses with R version 4.3.1.

### Systematic review and meta-analysis

We searched MEDLINE (PubMed) on January 21^st^, 2024, with the following search strategy: *(“SARS-CoV-2” OR “COVID-19” OR “COVID” OR “long COVID” or “post COVID-19 condition”) AND (“imatinib” OR “Gleevec” OR “Glivec”) AND (“RCT” OR “clinical trial” OR “randomized controlled trial” OR “random*”).* We additionally searched MedRxiv with the following search strategy: *“imatinib COVID”.* Two researchers independently screened search results for randomized trials reporting mortality for imatinib in hospitalized COVID-19, extracted contingency tables of overall mortality data at 30 days and 1 year, evaluated modified Cochrane risk of bias tool and assessed the certainty of evidence using the GRADE approach.^24^ We summarized the findings of our systematic review with a random-effects meta-analysis and a forest plot.

## Results

Between 6^th^ August 2021 and 20^th^ March 2023 we recruited 156 patients from 15 Finnish hospitals (103 patients recruited in 2021, 51 in 2022 and 2 in 2023, Supplementary Figure 1 and Supplementary Table 1). We randomized 83 patients (53%) to receive imatinib (with standard of care) and 73 (47%) to standard of care. One patient in the imatinib group did not receive imatinib, the only protocol violation of the trial.

We were able to retrieve overall mortality data for 156 (100%) patients at 30 days and 155 (99%) patients at 1 year (1 missing in standard of care). In the 1-year follow-up of those who survived we received responses from 62 (86%) patients in the imatinib group and 60 (91%) in the standard of care group. The overall response rate was 88% (122 responses) in patients still alive at 12-months post-randomisation (138 patients alive and followed for 1-year, 17 deceased, one did not yet complete 1-year follow-up). See Supplementary Figure 2 for a flowchart.

Table 1 describes patient characteristics, the relevance of these characteristics to in-hospital mortality, and their distribution between imatinib and standard of care group. Patient baseline characteristics were generally balanced between the two groups. The number of patients who received corticosteroids was 66 (80%) in the imatinib and 62 (85%) in the standard of care group. The median duration of treatment with imatinib was 6 days (IQR 3–9).

**Table 1.** Patient baseline characteristics by treatment assignment (imatinib or standard of care).

|  | Included in analyses |  |  |  | Treatment Assignment |  |
| --- | --- | --- | --- | --- | --- | --- |
| Characteristic | Entered study, n (%) | Mortality at 30 days*, n (%) | Mortality at 1 year, n* (%) |  | Imatinib, n (%) | Standard of care, n (%) |
| Number of patients | 156 (100) | 9 (6) | 17 (11) |  | 83 (100) | 73 (100) |
| Age (years) |  |  |  |  |  |  |
| <50 | 33 (21) | 0 | 2 (6) |  | 15 (18) | 18 (25) |
| 50-69 | 70 (45) | 0 | 2 (3) |  | 35 (42) | 35 (48) |
| ≥70 | 53 (34) | 9 (17) | 13 (25) |  | 33 (40) | 20 (27) |
| Sex |  |  |  |  |  |  |
| Men | 108 (69) | 7 (6) | 13 (12) |  | 56 (67) | 52 (71) |
| Women | 48 (31) | 2 (4) | 4 (9) |  | 27 (33) | 21 (29) |
| Major radiologic lung abnormality |  |  |  |  |  |  |
| No | 50 (32) | 2 (4) | 6 (12) |  | 27 (33) | 23 (32) |
| Yes | 99 (63) | 7 (7) | 10 (10) |  | 53 (64) | 46 (63) |
| Not imaged | 7 (4) | 0 | 1 (14) |  | 3 (4) | 4 (5) |
| Days to hospitalisation from start of symptoms |  |  |  |  |  |  |
| Less than 7 days | 68 (44) | 7 (10) | 11 (16) |  | 20 (24) | 17 (23) |
| 7-13 days | 82 (53) | 2 (2) | 6 (7) |  | 56 (67) | 51 (70) |
| 14 days or more | 6 (4) | 0 | 0 |  | 7 (8) | 5 (7) |
| Days in hospital at randomisation |  |  |  |  |  |  |
| 0 | 12 (8) | 0 | 0 |  | 9 (11) | 3 (4) |
| 1 | 69 (44) | 5 (7) | 11 (16) |  | 30 (36) | 39 (53) |
| ≥2 | 75 (48) | 4 (5) | 6 (8) |  | 44 (53) | 31 (42) |
| Location |  |  |  |  |  |  |
| University hospital | 103 (66) | 7 (7) | 11 (11) |  | 54 (65) | 49 (67) |
| Other hospital | 53 (34) | 2 (4) | 6 (11) |  | 29 (35) | 24 (33) |
| Current smoker | 16 (10) | 0 | 2 (13) |  | 9 (11) | 7 (10) |
| History of |  |  |  |  |  |  |
| Diabetes | 38 (24) | 2 (5) | 4 (11) |  | 21 (25) | 17 (23) |
| Heart disease | 43 (28) | 5 (12) | 7 (17) |  | 28 (34) | 15 (21) |
| Chronic liver disease | 1 (1) | 0 | 1 (100) |  | 1 (1) | 0 (0) |
| Chronic lung disease | 28 (18) | 4 (14) | 7 (25) |  | 15 (18) | 13 (18) |
| Asthma | 21 (13) | 0 | 2 (10) |  | 11 (13) | 10 (14) |
| Obesity (BMI 30 or more) | 71 (46) | 1 (1) | 4 (6) |  | 38 (46) | 33 (45) |
| COVID-19 severity at admission |  |  |  |  |  |  |
| No supplementary oxygen | 29 (19) | 1 (3) | 5 (18) |  | 16 (19) | 13 (18) |
| Low- or high flow oxygen | 118 (76) | 6 (5) | 10 (8) |  | 60 (72) | 58 (79) |
| Non-invasive ventilation | 9 (6) | 2 (22) | 2 (22) |  | 7 (8) | 2 (3) |
| Invasive ventilation | 0 | 0 | 0 |  | 0 | 0 |

\* Percentages are calculated as proportion of followed-up patients within each row.

### Hospital outcomes and 30-day mortality (short-term follow-up)

Figure 1A presents the cumulative incidence of overall mortality at 30 days. At 30 days, 6 patients had died in the imatinib group versus 3 in the group receiving standard of care. The adjusted HR for overall mortality at 30 days was 1.09 (95% CI 0.23–5.07) at 30 days (Supplementary Table 2 provides unadjusted HRs).

**Figure 1.**
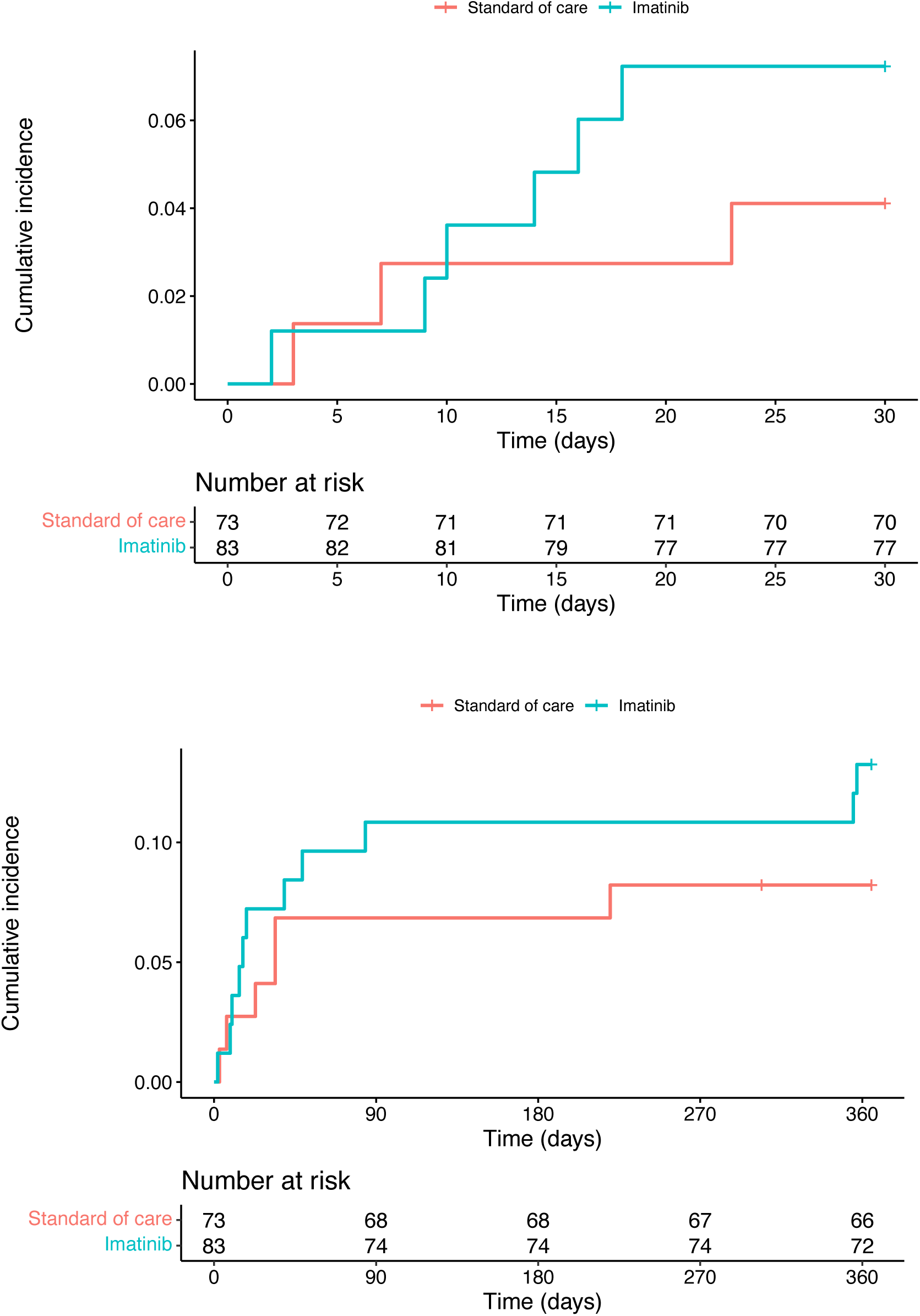
Kaplan-Meier curves of cumulative incidence of all-cause mortality in patients receiving either standard of care or imatinib evaluated at 30 days (Figure 1A) and 1 year (Figure 1B).

The mean duration of hospital stay was 11.1 days (standard deviation, SD 8.1 days) in the imatinib and 12.2 days (SD 13.6) in the standard of care group (absolute difference 1.1 days, 95% CI −2.6–4.6 days).

The number of patients with the need of invasive mechanical ventilation was 3 (4%) in the imatinib and 4 (5%) in the standard of care group (RR 0.66, 95% CI 0.15–2.85). Three patients with invasive ventilation additionally received extracorporeal membrane oxygenation (1 in imatinib, 2 in standard of care). The number of patients not receiving any supplementary oxygen was 16 (19%) in the imatinib and 11 (15%) in the standard of care group (RR 1.28, 95% CI 0.64–2.58). Supplementary Table 3 provides details on need of respiratory support during hospitalisation (no differences between groups).

Liver enzymes elevated in 5 (6%) patients in the imatinib and no patients in the standard of care group. These five patients discontinued imatinib without any drug-related consequences. One patient discontinued imatinib when thrombocytes dropped (from 92 to 40 E9/l) after one day of use. One patient discontinued imatinib (after 2 days of use) due to experience of nausea and diarrhoea, and another patient (after the first dosage) due to experience of mild dizziness and a strange sensation in the head.

### Long-term follow-up

Figure 1B presents 1-year cumulative incidence of overall mortality. At 1 year, 11 (14%) patients had died in the imatinib and 6 (8%) in the standard of care group. The adjusted HR was 1.34 (95% CI 0.46–3.88) at 1 year.

Table 2 and Figure 2 present patient important outcomes evaluated at 1-year post-randomisation. Results regarding having fully or largely recovered from their COVID-19 infection did not convincingly differ, and EQ-VAS scores were very similar.

**Figure 2.**
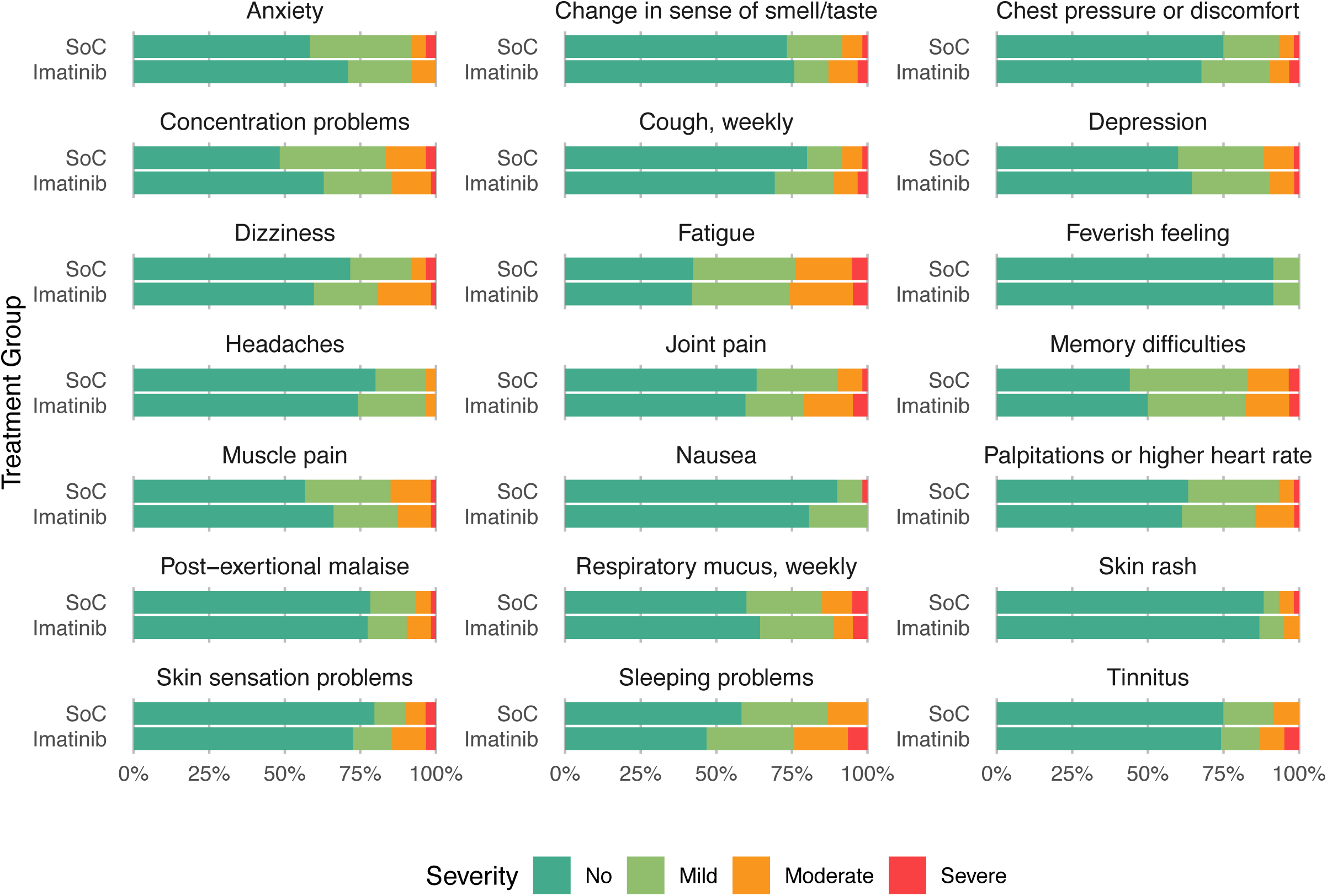
Outcomes possibly associated with long COVID evaluated at 12 months after hospital admission for patients receiving only standard of care (SoC) and standard of care plus imatinib.

**Table 2.** Effect of treatment group (imatinib or standard of care, SoC) on patient-important outcomes 1 year after hospitalisation due to COVID-19 infection.

| **Outcome** | **Imatinib,**<br>**n=62 (%)** | **Standard of**<br>**care, n=60 (%)** | **RR, 95% CI** |
| --- | --- | --- | --- |
| **How do you feel you have recovered from the COVID-19 infection you had a year ago?** |  |  | 0.91, 0.78–1.06 |
| Fully or largely (1–2) | 49 (79.0) | 53 (88.3) |  |
| About halfway recovered to not recovered at all (3–5) | 12 (19.4) | 7 (11.7) |  |
| **Exertional dyspnoea, mMRC dyspnoea scale** |  |  | 1.13, 0.67–1.89 |
| No to slight dyspnoea (mMRC 0–1) | 40 (65.6) | 41 (68.3) |  |
| At least a need to walk slower than usually (mMRC 2–4) | 21 (34.4) | 18 (30.0) |  |
| **Fatigue** |  |  | 0.95, 0.20–4.53 |
| No or slight fatigue (1–2) | 59 (95.2) | 56 (93.3) |  |
| Moderate or severe fatigue (3–4) | 3 (4.8) | 3 (5.0) |  |
| **Mobility, walking (EQ-5D-5L)** |  |  | 1.16, 0.54–2.48 |
| No or slight problems (1–2) | 50 (80.6) | 50 (83.3) |  |
| From moderate problems to unable to walk (3–5) | 12 (19.4) | 10 (16.7) |  |
| **Self-care, washing or dressing oneself (EQ-5D-5L)** |  |  | 0.97, 0.33–2.83 |
| No or slight problems (1–2) | 56 (90.3) | 54 (90.0) |  |
| From moderate problems to inability to wash or dress (3–5) | 6 (9.7) | 6 (10.0) |  |
| **Usual activities, e.g., work, study, housework, family or leisure activities (EQ-5D-5L)** |  |  | 0.83, 0.30–2.33 |
| No or slight problems (1–2) | 56 (90.3) | 53 (88.3) |  |
| From moderate problems to inability to do usual activities (3–5) | 6 (9.7) | 7 (11.7) |  |
| **Pain or discomfort (EQ-5D-5L)** |  |  | 0.82, 0.40–1.68 |
| No or slight pain (1–2) | 51 (82.3) | 47 (78.3) |  |
| From moderate to extreme pain (3–5) | 11 (17.7) | 13 (21.7) |  |
| **Anxiety or depression (EQ-5D-5L)** |  |  | 1.29, 0.30–5.52 |
| No or slight problems (1–2) | 58 (93.5) | 57 (95.0) |  |
| From moderate to extreme problems (3–5) | 4 (6.5) | 3 (5.0) |  |

Among the most common problems typically associated with long COVID observed in patients at the long-term follow-up were fatigue (24%), sleeping problems (19%) and memory difficulties (17%) (Figure 3). All differences proved easily attributable to chance.

**Figure 3.**
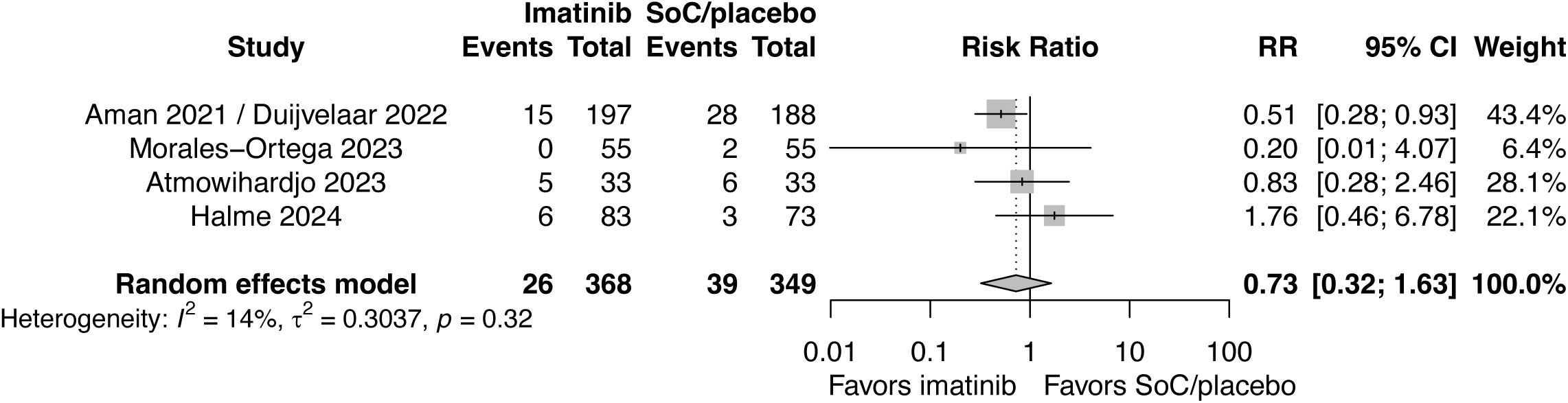
Random-effects meta-analysis of the effect of imatinib on 30-day overall mortality in randomized trials comparing imatinib to either standard of care or placebo in patients hospitalized for COVID-19.

### Systematic review and meta-analysis

Our search found three earlier randomised trials reporting on effect of imatinib on mortality (Supplementary Table 4). Figure 3 presents the results of the meta-analysis, including three earlier trials and our trial, including 732 patients. The risk ratio for the effect of imatinib treatment on overall mortality at 30 days was 0.73 (95% CI 0.32–1.63). We judged all studies low risk of bias for mortality (Supplementary Table 5). We rated down the evidence certainty (quality) due to very serious limitations in precision (confidence intervals including both appreciable benefit and appreciable harm). We therefore rated evidence certainty as low.

## Discussion

Solidarity Finland is the second largest randomised trial to report results of the effect of imatinib in patients hospitalized with COVID-19 and the only trial with long-term follow up of one year, including assessment of long COVID symptoms. We found no apparent short- or long-term benefits for imatinib in patients hospitalized for COVID-19 including survival, need for respiratory support or length of hospital stay at 30 days Similarly, we found no apparent benefit of imatinib on outcomes at 1 year at which time one in seven survivors reported that they had not fully recovered from their COVID-19 infection, one in four reported fatigue and one in five sleeping problems. A meta-analysis of three earlier trials and Solidarity Finland raises serious doubts regarding the benefit of imatinib in reducing mortality, improving recovery and preventing potential long COVID symptoms in patients hospitalized for COVID-19.

The major limitation of our study is the small sample size that leads to lower precision characterized by wide confidence intervals and higher risk of imbalances in baseline characteristics. To account for potential imbalances, we adjusted our analyses for baseline COVID-19 severity, age and sex which are likely prognostic factors for COVID-19 mortality. Although adjustment for these factors slightly decreased the hazard ratios, none of the adjusted nor unadjusted hazard ratios approached statistical significance.

Another limitation of our study may be the lack of blinding. In general, blinding is less important for objective than subjective outcomes (mortality vs symptoms). Lack of blinding is probably not an important issue for mortality in our trial. The finding of no important difference in the use of corticosteroids between imatinib and standard of care of arms suggests that lack of blinding did not result in important cointervention. In addition, importance of blinding on patient-important outcomes, such as recovery, long COVID or quality of life, is likely smaller in trials that examine symptoms at a longer duration from receipt of medication (one year in our case) rather than at the time of treatment of symptoms (such as long COVID treatment trials where symptoms are surveyed during treatment).

This study has several strengths. First, we performed a pragmatic, nationwide trial in 15 hospitals at university, central and city hospital levels. We used pragmatic study design with limited exclusion criteria to achieve broad patient population applicable to clinical practice representing hospitalized COVID-19 patients in Finland, including also immigrant groups. Second, we achieved a very high follow-up rate for both objective (mortality information for 98% at 1 year) and subjective outcomes (90% of survivors). To increase participation and avoid miscommunication and misunderstanding, we translated the questionnaire into nine languages and interpreters participated in phone interviews when necessary. Third, our multidisciplinary team of clinicians (representing eight different fields), methodologists, and patient partners created a questionnaire that focused on the most patient-relevant outcomes. To place our research within a broader context, we performed a systematic review of our own and three earlier trials studying the impact of imatinib on mortality in patients hospitalized for COVID-19. Our meta-analysis pooled data of all trials on mortality as well as assessing risk of bias and evidence certainty using the GRADE approach.

The first randomized trial on Imatinib for COVID - conducted in the Netherlands between March 2020 and January 2021 in non-vaccinated hospitalised patients–-was unable to meet its predefined primary outcome, defined as reduced time to discontinuation of ventilation and supplemental oxygen for more than 48 consecutive hours in patients with COVID-19 requiring supplemental oxygen. They found, however, an unadjusted HR of 0.51 (95% CI 0.27–0.95) for overall mortality and a significant difference in median duration of invasive mechanical ventilation (7 days in the imatinib vs 12 days in the placebo group, p=0.008) at 28 days follow-up. The result remained similar at 90 days follow-up (unadjusted HR for mortality 0.53, 95% CI 0.29–0.94).^25^

While the first trial was the largest (400 patients)^15,25^, our study had 156 patients, followed by one trial from Spain (110 patients)^17^, and another from the Netherlands (66 patients)^16^. Since the first trial, no other trial, including ours, has been able to show significant benefit of imatinib on mortality in patients hospitalized for COVID-19. Consequently, pooled results from trials raise serious doubts regarding the benefit of imatinib in reducing mortality when given to patients hospitalized for COVID-19 (Figure 3).

The first trial from the Netherlands recruited hospitalized COVID-19 patients requiring supplemental oxygen (to maintain a peripheral oxygen saturation of > 94%). Authors reported that imatinib particularly benefits patients with a severe course of COVID-19.^15^ They therefore conducted the subsequent trial that recruited invasively ventilated patients with moderate-to severe COVID-19 ARDS.^16^ The subsequent, smaller trial did not, however, find any impact of imatinib on mortality in patients with moderate-to severe COVID-19 ARDS.

A single-center trial from Spain was the first stage of a pick-the-winner trial between bariticinib and imatinib (with the third arm of standard of care).^17^ Authors recruited 55 patients to the imatinib arm (at 70 days follow-up, 2 patients died in standard of care, 2 in imatinib and none in the bariticinib arms). Our and the Spanish trial did not have any eligibility criteria regarding supplemental oxygen. In our trial, 19% had no supplementary oxygen, 76% had low-or high flow supplemental oxygen, 6% had non-invasive ventilation and none had invasive ventilation at hospital admission. Therefore, our (and the Spanish) trial had less sick patient population than the trials conducted in the Netherlands.

Solidarity Finland is the only imatinib trial with follow-up of mortality and recovery as well as quality of life and long COVID symptoms 1-year post-discharge. There are very few randomized trials reporting 1-year follow-up results following any treatment for COVID-19.^18,19^ We earlier reported our trial evaluating remdesivir in patients hospitalized for COVID-19, also a part of Solidarity Finland trials, using the same questionnaire.^18^ The international Solidarity trial found modest benefit for remdesivir in hospitalized COVID-19 patients.^26,27^ Although our Finnish in-hospital phase results were consistent with the global study, we were not able to find an effect on any long COVID outcomes.^28^

In a randomized trial of convalescent plasma (n=30) versus usual care (n=20) among German patients who survived severe COVID-19, authors compared quality of life at 1 year between. This small trial was unable to detect differences. As in our current imatinib and our earlier remdesivir trial, patients often reported substantial symptom burden at 1-year post-hospitalisation. Further studies with control groups without confirmed COVID-19 infection are needed to improve understanding of the long-term burden of disease.

In conclusion, we report the results of the Solidarity Finland trial evaluating the effect of imatinib on mortality and long-term outcomes in patients hospitalized for COVID-19. We found that imatinib conferred no treatment benefit on overall mortality evaluated at 30 days or 1 year. In addition, imatinib did not have an effect on long-term outcomes typically associated with COVID-19. However, 16% of respondents reported not having recovered from COVID-19 with fatigue, sleeping problems and memory difficulties among the most common symptoms. Our results are limited by our small sample size leading to inadequate statistical power in determining possibly small treatment effects. Further large pragmatic randomized trials with adequate follow-up are needed to rationalize drug treatment of COVID-19 as well as its associated long-term sequelae.

## Data Availability

The dataset generated during and analysed during the current study are not publicly available for data security. The corresponding author (K.A.O.T.) is the custodian of the long-term follow-up data and will provide access to de-identified and processed participant data for academic purposes within 2 months on request with the completion of a data access agreement.

## Acknowledgments

We thank professors Anssi Auvinen, Katri Kaukinen and Miia Turpeinen for serving as members of the data safety and monitoring board and Yvonne Peltonen for monitoring the trial. We thank Kaisa Harkman, Suzana Hentunen, Jenni Jouppila, Maiju Leppänen, Eveliina Muilu, Jenni Nykänen, Susanna Pieskä, Kalle Voutilainen and Terhi Wilppu for their expert guidance and dedicated support in pharmaceutical management. The World Health Organization (WHO) provided the study drug (imatinib) donated by Novartis. The study was funded by the Research Council of Finland (335527), Finnish Medical Foundation, Foundation of the Finnish Anti-Tuberculosis Association, Helsinki University Hospital State Research Funding (TYH2022330; TYH2023236), Paulo Foundation, Päivikki and Sakari Sohlberg Foundation, Research Foundation of the Pulmonary Diseases, Sigrid Jusélius Foundation, Tampere Tuberculosis Foundation and Tampere University Hospital State Research Funding (9AC085).

## Author contributions

J.R., O.P.O.N., and K.A.O.T. conceptualised the trial and contributed to its design. All authors were involved in the acquisition, analysis, and/or interpretation of data. A.L.E.H., J.R., N.P., G.H.G. and K.A.O.T. drafted the manuscript. All authors critically reviewed and approved the manuscript. A.L.E.H. and K.A.O.T. performed the statistical analyses. J.R. and K.A.O.T. obtained funding. K.A.O.T. supervised the study.

