## Supplementary Material for "Short- and long-term effects of imatinib in patients hospitalised for COVID-19 infection: A randomised controlled trial"

**Supplementary results**

**Figure 1. Patient recruitment chart in the Solidarity Finland imatinib trial**
The World Health Organization suspended the recruitment in the end of March 2023

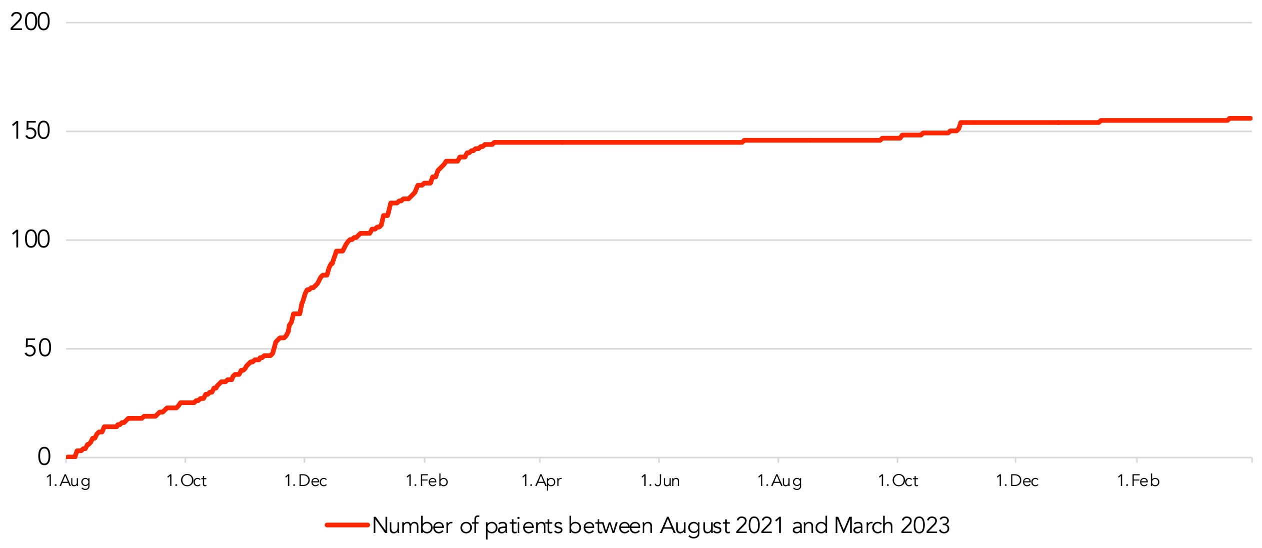

**Figure 2. Study flowchart**

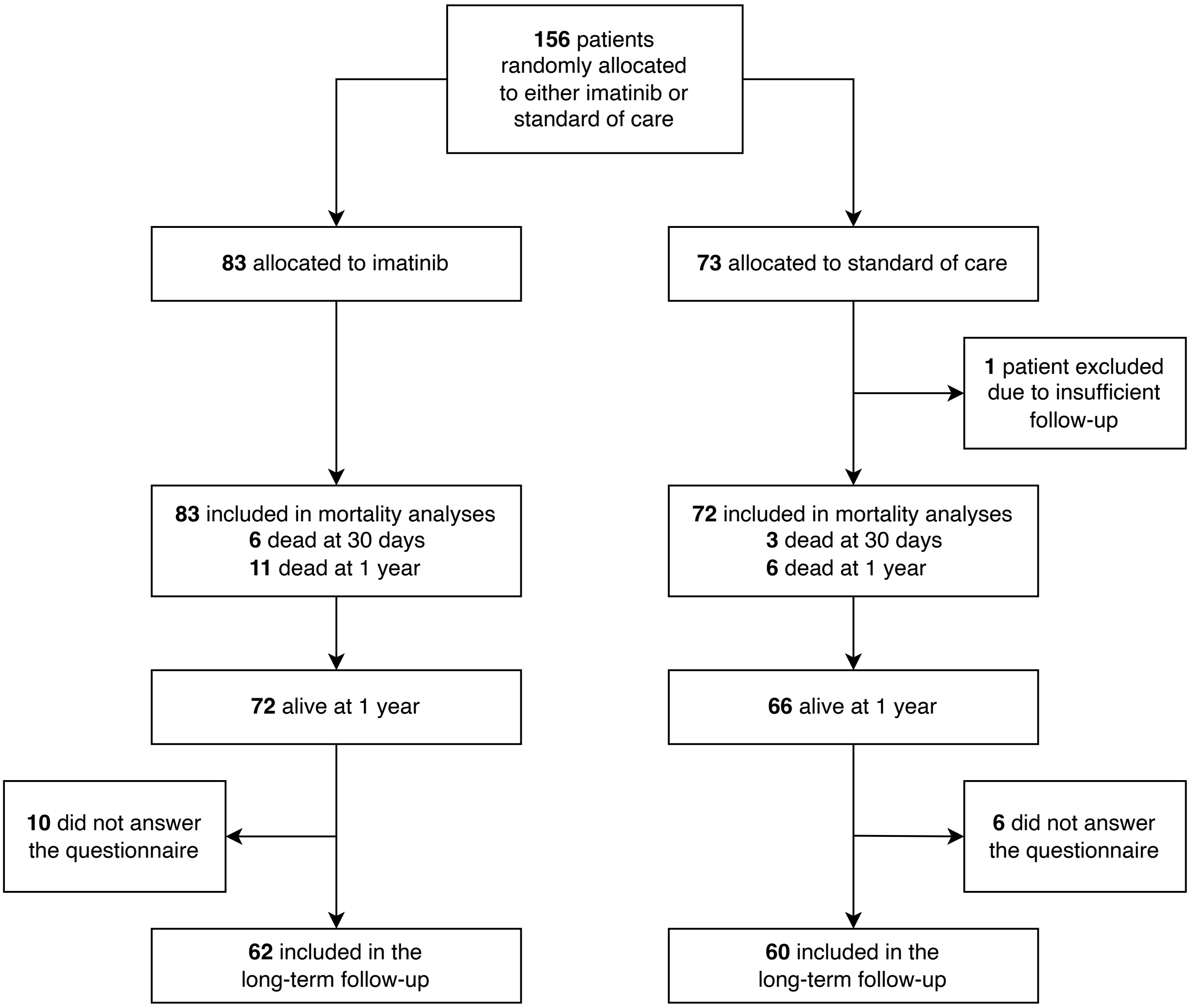

**Table 1. Number of recruited patients by hospital**

| Hospital | Recruited patients |
|---|---|
| Helsinki University Hospital (Meilahti, Jorvi, Peijas) | 40 |
| Tampere University Hospital (Keskussairaala, Hatanpää) | 38 |
| Kanta-Häme Central Hospital | 22 |
| Helsinki City Hospital (Laakso, Herttoniemi) | 22 |
| Oulu University Hospital | 20 |
| Hyvinkää Hospital | 5 |
| Kuopio University Hospital | 4 |
| Seinäjoki Central Hospital | 2 |
| Turku University Hospital | 1 |
| Mikkeli Central Hospital | 1 |
| Päijät-Häme Central Hospital | 1 |
| **Total** | **156** |

**Table 2. Unadjusted hazard ratios for mortality**

| Follow-up time | Unadjusted hazard ratio | 95% confidence interval |
|---|---|---|
| 30 days | 1.78 | 0.45–7.12 |
| 1 year | 1.64 | 0.61–4.44 |

**Table 3. Need of respiratory support during hospitalisation**

| | Imatinib (%) | Standard of care (%) | RR | 95% CI |
|---|---|---|---|---|
| No supplementary oxygen | 16 (19) | 11 (15) | 1.28 | 0.64–2.58 |
| Low-flow supplementary oxygen | 36 (43) | 41 (56) | 0.77 | 0.56–1.06 |
| High-flow supplementary oxygen | 9 (11) | 6 (8) | 1.32 | 0.49–3.53 |
| Non-invasive ventilation | 19 (23) | 11 (15) | 1.52 | 0.78–2.98 |
| Invasive ventilation | 2 (2) | 2 (3) | 0.88 | 0.13–6.09 |
| Extracorporeal membrane oxygenation | 1 (1) | 2 (3) | 0.44 | 0.04–4.75 |

**Table 4. Characteristics of the included studies (systematic review and meta-analysis)**

| Study | Country | No. of participants | Mean age | Male % | Imatinib treatment | Control arm | Primary outcome |
|---|---|---|---|---|---|---|---|
| Aman 2021 / Duijvelaar 2022 | Netherlands | 385 | 64* | 69 | 800 mg per oral on day 0, 400 mg once daily on days 1–9 | Placebo | Time to discontinuation of mechanical ventilation and supplementary oxygen for more than 48 consecutive hours |
| Morales-Ortega 2023 | Spain | 66 | 55 | 72 | 400 mg per oral once a day for 7 days | Open-label | Time to discharge or reduction of 2 points on an ordinal scale of clinical status |
| Atmowihardjo 2023 | Netherlands | 110 | 63 | 58 | 200 mg intravenously twice daily for a maximum of 7 days | Placebo | Change in extravascular lung water index between days 1 and 4 |
| Halme 2024 | Finland | 156 | 62 | 69 | 400 mg per oral daily until discharge or 14 days in hospital | Open-label | Overall mortality at 30 days and 1 year |

*Median value

**Table 5. Risk of bias in the included studies (systematic review and meta-analysis)**

| | Allocation sequence generation | Allocation concealment | Blinding* | Loss to follow-up | Selective reporting | Other problems | Overall risk of bias |
|---|---|---|---|---|---|---|---|
| Aman 2021 | + | + | + | + | + | + | Low |
| Atmowihardjo 2023 | + | + | + | + | + | + | Low |
| Morales-Ortega 2023 | + | + | + | + | + | + | Low |
| Halme 2024 | + | + | + | + | + | + | Low |

*We did not rate up risk of bias in open-label studies when assessing risk of bias for overall mortality.
